# A Randomised Crossover Trial: Exploring the Dose-Response effect Of Carbohydrate restriction on glycaemia in people with type 2 diabetes (D-ROC2)

**DOI:** 10.1101/2021.05.30.21258049

**Authors:** Ebaa Al Ozairi, Muhammad Abdul-Ghani, Nick Oliver, Brandon Whitcher, Reem Al Awadi, Abeer El Samad, Etab Taghadom, Jumana Al-Kandari, Nicola Guess

## Abstract

**Aims:** The role of carbohydrate restriction in the management of glycaemia in type 2 diabetes (T2D) has been a subject of immense debate and controversy partly due to low-carbohydrate trials being confounded by multiple factors including degree of calorie restriction, dietary protein content, and by no clear definition of a low-carbohydrate diet. The current study sought to provide insight into the relationship between carbohydrate restriction and glycaemia by testing the effect of varying doses of carbohydrate on continuous glucose concentrations within a range of intakes defined as low-carbohydrate while controlling for confounding factors.

**Methods:** This was a randomised crossover trial in participants with T2D testing 5 different 6-day eucaloric, isocaloric dietary treatments with varying carbohydrate contents (10%, 15%, 20%, 25%, and 30% kcal). Diets were kept isocaloric by exchanging %kcal from carbohydrate with predominantly unsaturated fat, keeping protein constant at 15% kcal. Daily self-weighing was employed to ensure participants maintained their weight throughout each treatment arm. Between dietary treatments, participants underwent a washout period of at least 7 days and were advised to maintain their habitual diet. Glycemic control was assessed using a continuous glucose monitoring device that was placed while the participant was on their normal diet, and was worn for the 6 days of each treatment.

**Results:** 12 participants completed the study. There were no differences in 24-hour and postprandial sensor glucose concentrations between the 30%kcal and 10%kcal doses (7.4 ± 1.1mmol/L vs 7.6 ± 1.4mmol/L (P=0.28) and 8.0 ± 1.4mmol/L vs 8.3 ± 1.3mmol/L (P=0.28) respectively). In our exploratory analyses we did not find any dose-response relationship between carbohydrate intake and glycaemia. A small amount of weight loss occurred in each treatment arm (range: 0.4 to 1.1kg over the 6 days) but adjusting for these differences did not influence the primary or secondary outcomes.

**Conclusions:** Modest changes in dietary carbohydrate content in the absence of weight loss while keeping dietary protein intake constant do not appear to influence glucose concentrations in people with T2D.

**Trial Registration:** ISRCTN 11067343.

## BACKGROUND

The impact of carbohydrate restriction on glycaemia has been a subject of controversy in the dietary management of diabetes. Despite numerous long-term randomized controlled trials and meta-analyses having been dedicated to the subject, firm conclusions remain elusive [1, 2].

A major factor in the interpretation of the trials has been the presence of significant confounding factors within and between the randomised control trials conducted to date. For example, even within interventions designed to provide a very low carbohydrate (<50g/day) diet the self-reported carbohydrate intake varied from 49g/day (13%kcal) [3] to 190g/day (47%kcal) by the end of the trial [4]. Definitions of “low-carbohydrate” dietary interventions included within meta-analyses have ranged from 10%kcal [5] to 45%kcal [6].

Regression analysis of low-carbohydrate trials suggests the hypoglycaemic effect of carbohydrate restriction might begin at intakes below 30%kcal [6] but this is based on very few trials. Conducting a dose-response analysis of the effect of carbohydrate intake on glycaemia might help define the content of low-carbohydrate diets based on the physiological effect of carbohydrate restriction rather than an arbitrary cut-off.

Second, weight loss also varies immensely within and between trials [5]. Since glucose reduces with calorie-restricted dietary interventions [7], the variability in weight loss within and between trials precludes any firm conclusions regarding whether carbohydrate restriction per se an independent contributor.

Third, in some, but not all, trials, carbohydrate is replaced by different fatty acid sources as well as protein, often resulting in an absolute increase in dietary protein intake. Protein is a potent stimulator of insulin secretion as well as other incretin hormones [8, 9] and neither the individual trials themselves, nor meta-analyses [5, 6] have adjusted for the hypoglycaemic effect of protein [10]. It is well accepted in the nutrition literature that the interpretation of macronutrient-specific effects can only be derived after careful consideration of pairwise, isocaloric substitutions across a range of doses, as has been done for lipoprotein outcomes.

Finally, trials have included people on a range of medications at baseline [5]. The limited data available suggests that carbohydrate restriction may be superior at helping people come off hypoglycaemic medications [3, 11], but this is has not been systematically accounted for in all trials, and likely underestimates the impact carbohydrate restriction has on glycaemia.

This research study aims to advance the field by examining the dose-response effect of carbohydrate restriction within the range of currently defined low-carbohydrate diets on glycaemia in people with T2D. To control for the major confounding factors, each dose will be tested with a eucaloric diet maintaining protein at 15%kcal, in participants not currently utilizing hypoglycaemic medications.

## RESEARCH DESIGN AND METHODS

### Research design

This randomized crossover trial compared the effects of five 6-day isocaloric dietary assignments differing in the proportion of kcal from carbohydrate (10%, 15%, 20%, 25%, and 30% of kcal) on 24-h glycaemia in individuals with T2D.

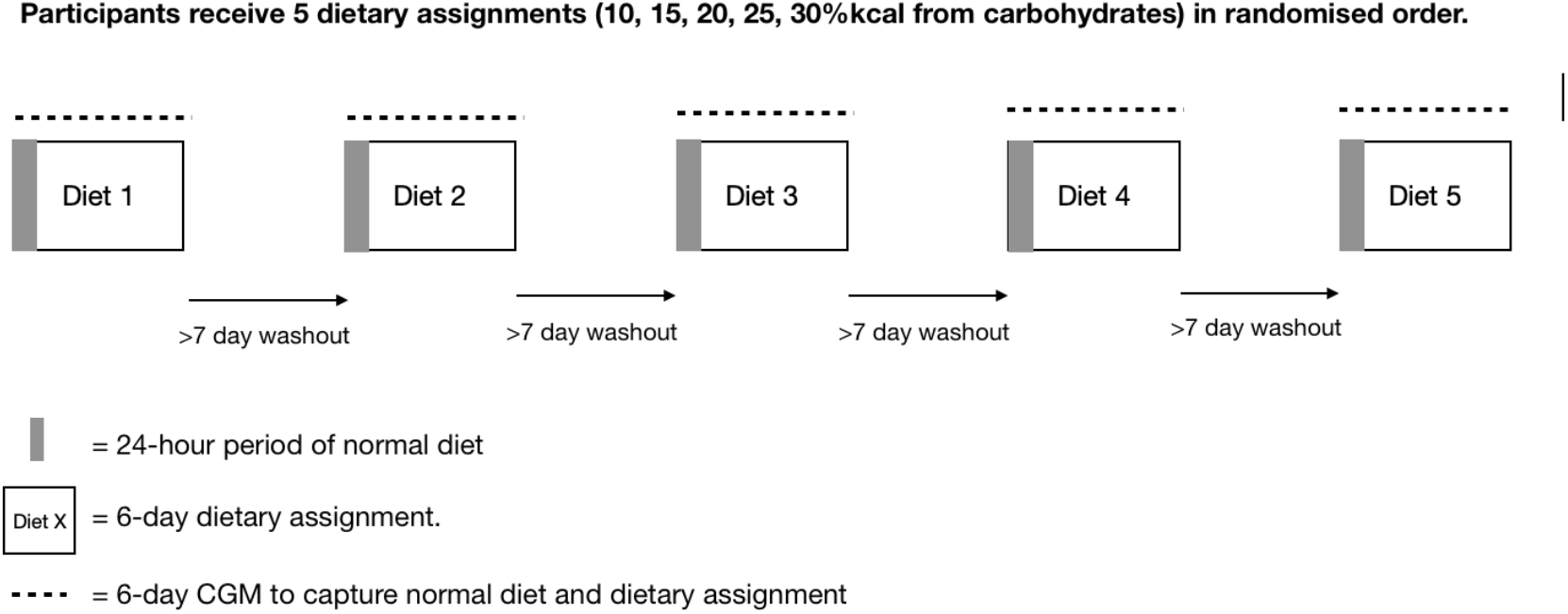

Prior to starting on each of their assigned “doses” of carbohydrate, participants were advised to continue their habitual diet. A continuous glucose monitoring device was placed 24-hours prior to the first meal of each dose to capture the change in glucose concentration between the normal diet and each carbohydrate restricted dietary assignment. A longer period of glucose concentration on the normal diet was not captured as the iPro2 sensor permits 7 days of wear. There is a physiological adjustment to carbohydrate restriction in the first 1-2 days [12]. During this 1-2 day period, glucose concentrations may be transiently reduced [12] which wouldn’t necessarily reflect the likely chronic effect of carbohydrate restriction on glycaemia. Therefore the 24-hour sensor glucose concentration and other glycemic outcomes for each “dose” was calculated from the final 4 days of each diet period.

Based on an expected mean 24-hour glucose of 8.9mmol/L [13] and aiming to power to see a difference of 0.9mmol/L (a 10% reduction) at alpha of 0.05 and 0.9 power we calculate that we needed 12 participants to see a difference between the highest and lowest dose. The additional between dose comparisons were exploratory.

#### Method

All recruitment and study visits were carried out at the Dasman Diabetes Institute in Kuwait between February 2019 and February 2020. A medical screening was conducted and reviewed by a medical doctor to ensure that participants were healthy and suitable for inclusion.

#### Inclusion criteria

Males and postmenopausal females aged 40–60 years who had a body mass index (BMI) of 25–40 kg/m^2^, reported a stable weight for 3 months prior to study commencement, were able to provide informed consent, and were willing to follow the diets described during the telephone screening were included. T2D had to be diagnosed within the last 3 years and controlled through diet or metformin, which was withdrawn 2 weeks prior to study initiation. Participants who had type 1 or monogenic diabetes, were on any medication for T2D other than metformin, and had kidney disease, liver disease, hematologic abnormalities, congestive heart failure, untreated thyroid disease cancer, and any other debilitating disease were excluded.

#### Dietary Intervention

This study compared 5 different levels of carbohydrate restriction which were within a range of intakes used within “low-carbohydrate” trials [5]. The amount of carbohydrate in the diet was based on the “biologically available carbohydrate” [14]. The lowest level of intake was set at 10%kcal, based on this being the suggested cut-off for a very-low-carbohydrate diet [5, 15]. The highest level of intake was set at 30%kcal to capture the degree of carbohydrate restriction at which we anticipated the glucose lowering impact might begin [6]. The proposed definition of low-carbohydrate diets is <26%kcal [15] so using 30%kcal would allow us to examine the impact of carbohydrate restriction at the levels of intake which might reasonably be considered “low-carbohydrate”. We defined the degree of carbohydrate restriction by the percentage of kcal to control for the differing energy requirements amongst our participants. Protein was kept at 15%kcal.

To encourage participants to follow their assigned dietary protocol, all meals were provided with clear instructions not to consume any other foods or calorific or carbohydrate-containing beverages. The National Institutes of Health Body Weight Planner [16] was used to determine the calorie intake required to maintain each participant’s body weight accounting for carbohydrate intake, and inputting individual physical activity level (PAL) based on self-reported usual activity. The model was not adjusted for sodium intake, as participants were allowed to use salt and pepper to flavour the meals. Meal plans were designed and assessed by NG to ensure that they corresponded with the macronutrient requirements of the study and were prepared and delivered by commercial medical meal providers (Basic, Kuwait City; DietFix, Kuwait City). Foods in Kuwait include local and important products. The Nutritics US and Arabic food databases was used to design the menus and where necessary the manufacturer was contacted to determine the macronutrient content of the foods. The macronutrient content of the diets at each level of intake is shown in (Table 1). To ensure that individual and collective meal plans were as similar as possible, the amount but not the type of food was adjusted first when planning the menus, after which small amounts of additional foods, such as oil or dates, were added to ensure the meals satisfied the macronutrient requirements. This ensured that any differences in the amount of fibre and sugar were minimised as much as possible. An example of how the amount of each food in the meal plans was altered to meet the macronutrient requirements of the dietary assignment is presented in supplementary data (Table S1). Although coffee and tea were allowed, participants were instructed not to change the usual amount consumed during the trial.

**Table 1:** Intake of macronutrients on each dietary assignment period. CHO = carbohydrate; SFA = saturated fat; UFA = unsaturated fat. * Sugar included glucose, sucrose, maltose and lactose.

|  | **10%** | **15%** | **20%** | **25%** | **30%** |
| --- | --- | --- | --- | --- | --- |
| **CHO (g/day)** | 48 to 93g | 71 to 139g | 95 to 185g | 119 to 231g | 143 to 278g |
| **CHO (%kcal)** | 10% | 15% | 20% | 25% | 30% |
| **Sugar\* (% total CHO)** | 36% | 33% | 31% | 28% | 27% |
| **Protein (g/day)** | 71 to 139g | 71 to 139g | 71 to 139g | 71 to 139g | 71 to 139g |
| **Protein (%kcal)** | 15% | 15% | 15% | 15% | 15% |
| **Fat (g/day)** | 158 to 308g | 148 to 288g | 137 to 267g | 127 to 247g | 116 to 226g |
| **Fat (%kcal)** | 75% | 70% | 65% | 60% | 55% |
| **SFA (%kcal)** | 25% | 24% | 22% | 21% | 19% |
| **SFA (%total fat)** | 35% | 34% | 33% | 35% | 35% |
| **SFA:UFA** | 1:3 | 1:3.2 | 1:3.6 | 1:3.8 | 1:4.2 |
| **Fibre (g/day)** | 15 to 26g | 16 to 27g | 18 to 28g | 20 to 31g | 21 to 35g |

Throughout each 7-day period during which the continuous glucose monitoring (CGM) was worn, participants were contacted at least once a day via text message or phone to ensure they had taken glucometer measures to calibrate the CGM, weighed themselves and were consuming their meals as required. Participants were provided with Bluetooth-enabled weighing scales with which to weigh themselves daily and ensure neither weight loss not gain. If a participant gained or lost 0.5 kg on two consecutive study days, they were advised to reduce or increase their intake while keeping the macronutrient content of the diet constant. Participants were advised not to change their usual physical activity habits and a pedometer (Nakosite BPED2433, Nakosite USA Ltd) was used to quantify the participants’ daily steps throughout each 7-day diet period.

There was a wash-out period of at least 7 days between each dietary assignment during which participants were asked to return to their habitual diet prior to entry into the study.

#### Continuous Glucose Monitoring

24-hours before participants were due to receive the first meal of each dietary assignment, a Medtronic iPro2 (Medtronic, Northridge, CA) blinded continuous glucose monitor (CGM) was placed on the anterior abdominal wall of each participant. Participants were then instructed to measure their capillary glucose concentrations at least twice daily using a provided glucometer (Accu-chek®) to calibrate the CGM.

On the last day of each dietary assignment, the iPro2 CGM sensor was removed, the data was downloaded, and the daily steps from the pedometer were recorded.

The order in which individuals were assigned in a 1:1 ratio to each level of carbohydrate intake was generated randomly using randomiser.org. N.G. carried out the randomisation, and informed the research assistants. Due to the nature of the study, neither the research staff, the participants were blinded to their allocation as they would be able to tell which week had more starchy foods. The primary and secondary data analyses were blinded.

### Outcomes

All glycaemic outcomes were calculated from the final 4 days of each dietary assignment. The primary outcome was the 24-h sensor glucose concentrations. The 24-h sensor glucose was defined as the mean of all interstitial glucose values measured using the CGM device every 5 min. Secondary outcomes included fasting sensor glucose and postprandial sensor glucose concentrations [17]. Fasting glucose was calculated as the mean of all interstitial glucose values measured using the CGM device every 5 min for the 2-h prior to breakfast. Postprandial glucose concentrations were calculated as the mean of all interstitial glucose values measured using the CGM device every 5 min for the 2-h postprandial period. Time zero was the meal time (whether breakfast, lunch, or dinner) recorded by the participant using the Accu-chek® glucometer. Other standardised measures of glycemic variability were also calculated including mean amplitude glucose excursions (MAGE), X continuous overlapping net glycemic action (CONGA) and the high blood glucose index (HBGI) and low blood glucose index (LBGI).

### Statistical analysis

The difference in sensor glucose concentrations between the highest and lowest dose was determined using a paired *t* test. A linear mixed-effects model was used to explore the relationship between dose and glycemic outcomes outcomes. The dose response was assumed to be linear for the primary and secondary outcomes. Fixed effects in the model included dose, age, biological sex, whether or not subjects were from Kuwait, difference in body weight during each dose, and average number of daily steps performed. Four missing values were observed in the average number of daily steps performed and were imputed using the average across all dose levels for the subject. Subjects were considered a random effect, and their impact on the intercept and dose response was investigated. Analysis of covariance was used to compare differences between doses while accounting for weight change and steps per day after testing for normality using the Kolmogorov–Smirnov test. Statistical analyses were performed using SPSS (version 26) and R (version 3.6.3), with P ≤ 0.05 indicating statistical significance.

## RESULTS

Among the 26 individuals screened for the study, 19 satisfied the inclusion criteria and agreed to participate. Four other individuals declined to participate after randomization but before the trial started. Among the 15 individuals who participated in the study, 12 completed it (Flowchart is Figure S1 in supplementary data). Table 2 summarizes the baseline patient characteristics.

**Table 2:** Demographic information for the n=15 people who started the study. *Median (IQR) †Ratio

| Characteristic | Mean ± SD |
| --- | --- |
| Age (years) | 54 (47 to 56)* |
| Gender (M:F) | 9:6<sup>2</sup> |
| Weight (kg) | 84.2 ± 13.4 |
| BMI (kg/m<sup>2</sup>) | 30.8 ± 4.3 |
| FPG (mmol/L) | 8.1 (7.3 to 9.0)† |
| HbA1c (%) | 6.6 ± 0.6.<br>(49 ± 0.9 mmol/mol) |
| Years diagnosed with T2D |  |
| 0 | n=4 |
| 1 | n=8 |
| 2 | n=1 |
| 4 | n=2 |
| Metformin Use |  |
| Yes | n=10 |
| No | n=5 |

### Glycaemic outcomes

The unadjusted mean and standard deviation of 24-hour glucose, postprandial glucose, fasting glucose and glycaemic variability measures are shown in Table 3. There were no differences difference between the 10%kcal and 30%kcal dose for 24-hour glucose and post-prandial glucose sensor concentrations: 7.4 ± 1.1mmol/L vs 7.6 ± 1.4mmol/L (P=0.28) and 8.0 ± 1.4mmol/L vs 8.3 ± 1.3mmol/L (P=0.28) respectively. Fasting glucose was also not different between the 10%kcal and 30% kcal dose: 7.0 ± 1.3 vs 7.1 ± 1.6 (P=0.59).

**Table 3:** Sensor glucose parameters over the final four days of each dietary assignment period (CONGA (continuous overall net glycaemic action); MAGE (mean amplitude of glucose excursions); LBGI (low blood glucose index); HBGI (high blood glucose index)). Mean weight change over the 7-day duration of each dosing period, and mean steps per day during each carbohydrate dosing period. The figures are mean ± SD. *Difference between groups, P=0.086); †Difference between groups, P=0.98). PP = post prandial. The P value shows the significance level between the 10% and 30% doses.

|  | 10% | 15% | 20% | 25% | 30% | P value |
| --- | --- | --- | --- | --- | --- | --- |
| 24-h glucose (normal diet) | 7.3 ± 1.2 | 7.4 ± 1.6 | 8.0 ± 1.4 | 7.7 ± 1.3 | 7.5 ± 1.5 | 0.33 |
| 24-h glucose (assigned diet) | 7.4 ± 1.1 | 7.1 ± 1.2 | 7.6 ± 1.4 | 7.6 ± 1.2 | 7.6 ± 1.3 | 0.28 |
| Fasting glucose (assigned diet) | 7.0 ± 1.3 | 6.8 ± 1.1 | 7.1 ± 5.6 | 7.2 ± 1.2 | 7.1 ± 1.6 | 0.59 |
| PP glucose (assigned diet) | 8.1 ± 1.5 | 7.9 ± 1.4 | 8.3 ± 1.6 | 8.5 ± 1.5 | 8.5 ± 1.4 | 0.28 |
| CONGA | 1.5 ± 0.6 | 1.7. ± 0.6 | 1.6 ± 0.5 | 1.9 ± 0.6 | 1.6. ± 0.4 | 0.85 |
| MAGE | 3.6. ± 1.5 | 4.0. ± 1.6 | 4.4. ± 1.1 | 4.4 ± 1.6 | 3.6. ± 1.2 | 0.94 |
| LBGI | 0.3. ± 0.4 | 0.5. ± 0.7 | 0.2. ± 0.4 | 0.4. ± 0.5 | 0.3. ± 0.5 | 0.95 |
| HBGI | 2.3. ± 2.3 | 2.6. ± 2.0 | 2.8 ± 2.7 | 3.2. ± 2.3 | 2.8. ± 2.6 | 0.28 |
| Weight change (kg)* | -0.51 ± 0.83 | -0.58 ± 0.67 | -1.14 ± 0.88 | -0.38 ± 0.4 | -0.9 ± 0.9 | 0.29 |
| Mean steps per day† | 5505 ± 2406 | 5416 ± 3746 | 4871 ± 2367 | 4918 ± 2947 | 5282 ± 2292 | 0.64 |

We then explored the dose-response relationships between carbohydrate dose and glycaemia using mixed-effect modelling. There was no relationship between dose of carbohydrate and the mean 24-h glucose concentration (Figure 1a), postprandial glucose concentration (Figure 1b), or any of the secondary glycaemic outcomes (P > 0.05). Including random slopes for the dose response did not improve the any of the models according to a likelihood ratio test (P > 0.05). Age and biological sex were identified as parameters with a significant effect on mean 24-h glucose concentration (P < 0.05). Age had a significant effect on postprandial glucose concentration (P < 0.05), weight change had a significant effect on MAGE (P < 0.05), and gender had a significant effect on both log (LBGI) and HBGI (P < 0.05). The individual postprandial glucose excursions for breakfast, lunch, and dinner for each of the doses are illustrated in supplementary data (Figure S1). Parameter estimates from the linear mixed-effects models are shown in supplementary data (table S3).

**Figure 1:**
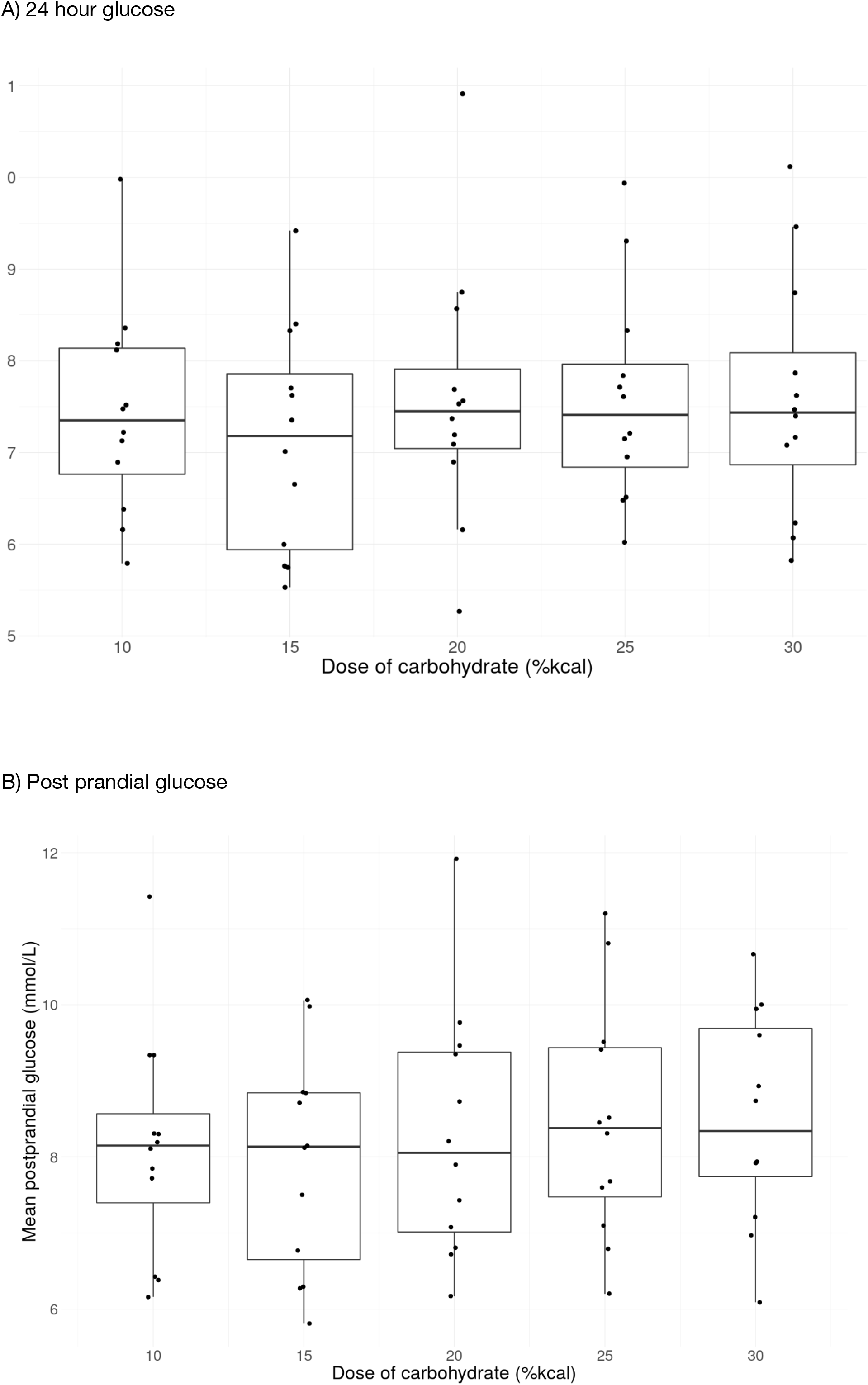
Mean sensor a) 24-hour and b) post-prandial glucose concentrations for each of the carbohydrate doses over the last s4 days of each dosing period. N=12 for all observations. Each black filled dot represents one participant. The boxplots show the median and IQR. The whiskers represent >1.5 times the IQR from the median. The 24-hour sensor glucose concentration was determined by calculating the mean across all the interstitial glucose measure taken once every 5 minutes over the entire time the participant wore the device starting from the time from the very first assigned meal. The postprandial glucose concentration was determined by calculating the mean across all the interstitial glucose measures taken once every 5 minutes for the 120 minute period after every breakfast, lunch and dinner for each of the meals.

Finally, we explored the difference in 24-hour glucose concentrations during the normal diet and on each of the doses. There were no differences in blood glucose concentration between the normal diet and each of the doses (Table 3).

### Weight change

Although both diets were isocaloric, a small decrease in body weight was observed during each low carbohydrate diet, which reached within-group significance with a carbohydrate dose of 20% (78.2 ± 9.6 to 76.8 ± 9.3 kg; P = 0.001) and 30% (77.3 ± 7.3 to 76.2 ± 7.1 kg; P < 0.001). However, no significant differences in weight change was observed between the dietary assignments (Table 3).

### Physical activity

Mean daily steps ranged from ∼4800 to ∼5500 steps per day. No significant differences in steps per day were found between the dietary assignments (Table 3).

## DISCUSSION

In this study, we compared different levels of carbohydrate intake within isocaloric dietary assignments on glycaemia while maintaining protein. We did not find any differences in 24-hour, post-prandial or fasting sensor glucose concentrations between the 30%kcal and 10%kcal doses in this study. In our exploratory analyses we did not find a relationship between carbohydrate intake and glycaemia within the range of intakes. We think there are a number of points that other researchers can take from our work.

Firstly, we selected the levels of carbohydrate intake that we examined based on proposed, and widely used definitions of very low and low carbohydrate diets. Large differences in carbohydrate intake lower glucose concentrations in healthy individuals ((70%kcal to 10%kcal) KH study out 21st Jan) and reduce glycogenolysis in T2D (from 86%kcal to 6%kcal significantly) [18], when controlling for weight and protein. However, we wanted to exam the effect on glycaemia of more modest changes in carbohydrate intake. We did not observe any difference in glycaemia between the dietary assignments or when switching from the normal diet to any of the dietary assignments.

The calorie intake in this study required to maintain body weight ranged from 1900kcal to 3700kcal. At 1900kcal, increasing the %kcal from carbohydrate from 10%kcal to 30%kcal results in an increase in absolute intakes of carbohydrate from 48g/day to 142g/day. At 3700kcal the absolute grammes of carbohydrate increases from 93g/day to 278g/day. We find it surprising that we did not observe a difference in glycaemia even with a difference in carbohydrate intake as large as 185g/day. Our study was powered to detect a difference of 0.9mmol/L between the highest and lowest doses, which represents a 10% reduction in 24-hour glycaemia assuming a mean 24-jour glucose of 8.9mmol/L. We considered this to be a clinically relevant difference in glycamia. Our study was not powered to detect smaller differences in glucose concentration between dietary assignments, and therefore it is possible that a larger sample size might have been able to detect smaller differences in blood glucose between the doses of carbohydrate used in this study. Nevertheless, the fact that a change in absolute carbohydrate intake of 185g/day results in a change in glycaemia of < 0.9mmol/L suggests that very large differences in carbohydrate intake might be needed for any clinically relevant change in glucose concentration in T2D.

It is possible that our lowest carbohydrate dose did not go low enough. The lowest absolute intake in this study was 48g/day in the 10%kcal group for a 1900kcal intake. The range of absolute intake at the 10%kcal level was 48g/day to 93g/day across the cohort of 12. There is growing evidence that ketone bodies themselves independently lower glucose [19-21] at least partly by reducing hepatic glucose output [19] and long-term randomised [22] and open-label trials [23] which have used nutritional ketosis (>0.5mmol) as a therapeutic goal and a measure of compliance suggest that carbohydrate restriction in the context of significant weight loss and sufficient to induce ketosis may produce large reductions in both HbA1c and diabetes medication use. The degree of carbohydrate restriction required to induce ketosis is unclear and depends on a variety of factors, but has been suggested to be in the 20-30g/day range. It’s therefore possible that had we tested a level of intake at 5%kcal we would have seen significant reductions in glycaemia. Unfortunately we did not measure capillary or plasma ketones in this study which would have provided insight into this question.

A month-long inpatient study by Garg et al [24] compared a high carbohydrate diet (60%kcal) to a low carbohydrate diet (35%kcal), and found a significant reduction in mean glucose level and insulin dose in the low-carbohydrate group. In a separate study by the same group [25], there were significant reductions in post-prandial glucose following a 6-week low-carbohydrate diet (40%kcal) compared to the high-carbohydrate diet (55%kcal). Protein was kept at 15%kcal in both studies. However, a difference between their studies and ours is that they used a very high monounsaturated fat (25%kcal in [25] and 33%kcal in [24]) and low saturated fat content (10%kcal) in both low-carbohydrate groups. The monounsaturated fat content of their high-carbohydrate groups was 9%kcal [24] and 10%kcal [25]. Large differences in monounsaturated fat intake can influence insulin sensitivity [26] and lower glucose in T2D [27]. Replacing saturated fat sources with polyunsaturated or monounsaturated fat sources may also influence insulin secretion [28], and insulin sensitivity [26, 28, 29]. In this study, while the percentage of total fat kcal from saturated fat remained constant, the proportion of total kcal from saturated fat and consequently the ratio of saturated to unsaturated fat changed. This could have influenced our results. In Garg [25] there was an increase the saturated to unsaturated fat ratio of 1:3 to 1:4.5 which is comparable to the difference between the lowest and highest carbohydrate group in this study. For the time being it therefore it seems sensible to recommend the consumption of unsaturated fat to replace carbohydrate in people with T2D, especially given the protective effect of predominantly unsaturated fat foods on blood lipids related to CVD risk.

We kept each time period to 7 days. The effect of carbohydrate restriction (presumably, if the restriction is large enough) on glycaemia occurs with the first meal [30-32], and appears to be sustained at least in the medium term provided that the diet is followed [30]. So we expected to be able to see any effect of the carb restriction within a 7 day period. The shorter time period also allowed us to be as controlled as possible while minimising the burden on the participants. Since pronounced metabolic adaptations have been reported in the first 2-3 days of carbohydrate restriction [12, 33], including a transient reduction in blood glucose concentration [12] our outcome measurements were based on the final 3-4 days of each dietary period only. Nevertheless, it possible that there are other adaptations to carbohydrate restriction with may take longer [34] which we were not able to observe in the short length of time we captured.

Although T2D is often characterized as a condition of hyperinsulinemia, the postprandial insulin response to an increase in glucose concentration remains deficient. In contrast, beta-cells in individuals with T2D may maintain a relatively greater insulinogenic response to protein [8]. This often ignored discrepancy may explain why studies that only modestly reduce carbohydrate content while increasing protein content in individuals with T2D do indeed show marked reductions in glycaemia [10, 35, 36]: a robust postprandial insulin response can supress gluconeogenesis and hepatic glucose output, while promoting insulin-mediated glucose uptake [37, 38]. Although an increase in dietary protein has been associated with an increase in plasma glucagon [36], this is not sufficient to cause any meaningful increase in the rate glucose appearance in T2D [39]. This study was not designed to test the hypothesis that a high-protein, low-carbohydrate diet could lower glycaemia compared to a low-protein, low-carbohydrate diet. However, we do think that based on our current results, and the growing evidence of a hypoglycaemic effect of protein in T2D [9, 10, 35], the research community should clearly consider and account for the amount of protein in a nominally “low-carbohydrate” diet.

This study aimed for participants to maintain their body weight throughout each dietary assignment period in order to isolate the effect of carbohydrate restriction independent of a kcal deficit. Therefore, our findings cannot be extrapolated to assess the utility of low-carbohydrate within clinical practice for the management of T2D. Weight loss is a predominant driver of glyceaemic reduction in T2D [7], and low-carbohydrate diets are recognised as an effective way for many people to manage their weight [40].

This study has a number of strengths. We designed 420 separate menus for the 12 participants in order to control for the major confounding factors including weight loss, protein and saturated fat intake. Dose-response studies can permit a better understanding of the impact of food or nutrients on human metabolism [41] and to our knowledge, this is the first study to test a range of levels of carbohydrate intake on glycaemia in T2D. All meals were delivered to the participants daily, and we had daily contact with the participants to encourage them to weight themselves, consume their meals, and avoid any additional caloric items. Nevertheless, this was not a ward-based study and as such we could not observe the participants at all times. Therefore it’s possible that people did not stick to the dietary assignments. This remains the most important limitation, and it’s important to acknowledge only ward-based studied can truly test physiological efficacy. We do note that an advantage of CGM is that the glycemic excursion readouts can partly serve as a measure of compliance, especially because the consumption of a high-carbohydrate food during periods of carbohydrate restriction results in an exaggerated glycemic response. We did not see any evidence from the CGM read-outs that participants were consuming high-carbohydrate foods outside of the prescribed diets but we cannot know this for sure. Nevertheless, weight loss did occur in each of the groups, reaching clinical significance in the 20%kcal and 30%kcal ranges. This could indicate that either our estimates of the kcal required to maintain body weight were incorrect, or participants were not consuming all food provided to them. Nevertheless we do not think this meaningfully changed the outcomes, as adjusting for weight loss did not alter our primary findings.

A limitation of our study design is that we did not provide controlled diets during a run-in and washout period,, and did not assess dietary intake for the 24-hour period prior to starting each assigned dose. We designed the study on this basis because we expected to see a difference in glycaemia between the normal high-carbohydrate Kuwaiti diet (average intake 60%kcal from carbohydrates) and the carb-restricted doses - and were interested in the differences between the dietary assignments. This limitation means that we cannot within this study assess with any confidence whether larger differences in carbohydrate intake would have resulted in changes in glycaemia as it’s possible participants had already reduced their carbohydrate intake during the 24-hour period prior to commencing each dietary assignment.

We think this is unlikely given the Kuwaiti culinary culture and usual very high refined carb intake, though it remains possible. Finally, we tried to modify the amount of food before modifying the type of food to meet the macronutrient requirements of the diet to minimise differences in the type of carbohydrate, including fibre and sugar, but we did not succeed entirely. The amount of total sugar as a proportion of carbohydrate varied from 27% in the highest carbohydrate to 36% in the lowest carbohydrate group, and there was a difference of 5-10g of fibre per day between the lowest and highest carbohydrate groups. Much larger differences in sugar (whether sucrose or glucose)[42] are needed to see differences in glucose concentrations but it is possible that the difference in dietary fibre could have influenced our results [43]. The glycemic index of many of the Kuwaiti food products we used is unknown so we were unable to determine differences in glycemic index between the diets. However, we expect that since we manipulated the amount of foods not the type between groups that any difference in the glycaemic index between the diets would be very small. Since the glycaemic load is determined by both amount and the glycaemic index of foods, and since large differences in the glycaemic index [44] are needed to alter insulin sensitivity and glucose concentrations so we do not think this would have influenced our results.

## Conclusion

Modest short-term changes in carbohydrate intake do not appear to influence glycaemia in people with T2D when body weight and protein intake are maintained.

## Data Availability

The datasets during and/or analysed during the current study available from the corresponding author on reasonable request.

## List of abbreviations

CGM: continuous glucose monitoring
CONGA: continuous overlapping net glycemic action
HBGI: high blood glucose index
LBGI: low blood glucose index
MAGE: mean amplitude glucose excursions
PAL: physical activity level
T2D: type 2 diabetes.

## Declarations

Ethics approval and consent to participate

The trial was approved by the Dasman Diabetes Institute Research Ethics Committee (ref: RA HM-2018-04) and prospectively registered with the International Standard Randomised Controlled Trial Number (11067343).

## Authors’ contributions

N.G., E.A. and M.A. designed the study. N.O. provided specific input into the glycaemic outcomes in the planning of the study. R.A., A.S., E.T. and J.A. carried out recruitment, all study visits and data collection. N.G. oversaw the day-to-day management of the study. B.W. carried out the primary and secondary statistical analyses. N.G. wrote the manuscript with input and edits from N.O., E.A., M.A. and B.W. All authors reviewed and approved the final manuscript.

## Acknowledgements

We are grateful for the staff at Dasman Diabetes Institute for their support with the study. Finally, we are most grateful to the participants themselves who volunteered to take part in this study.

