## Supplementary Data for "A Randomised Crossover Trial: Exploring the Dose-Response effect Of Carbohydrate restriction on glycaemia in people with type 2 diabetes (D-ROC2)"

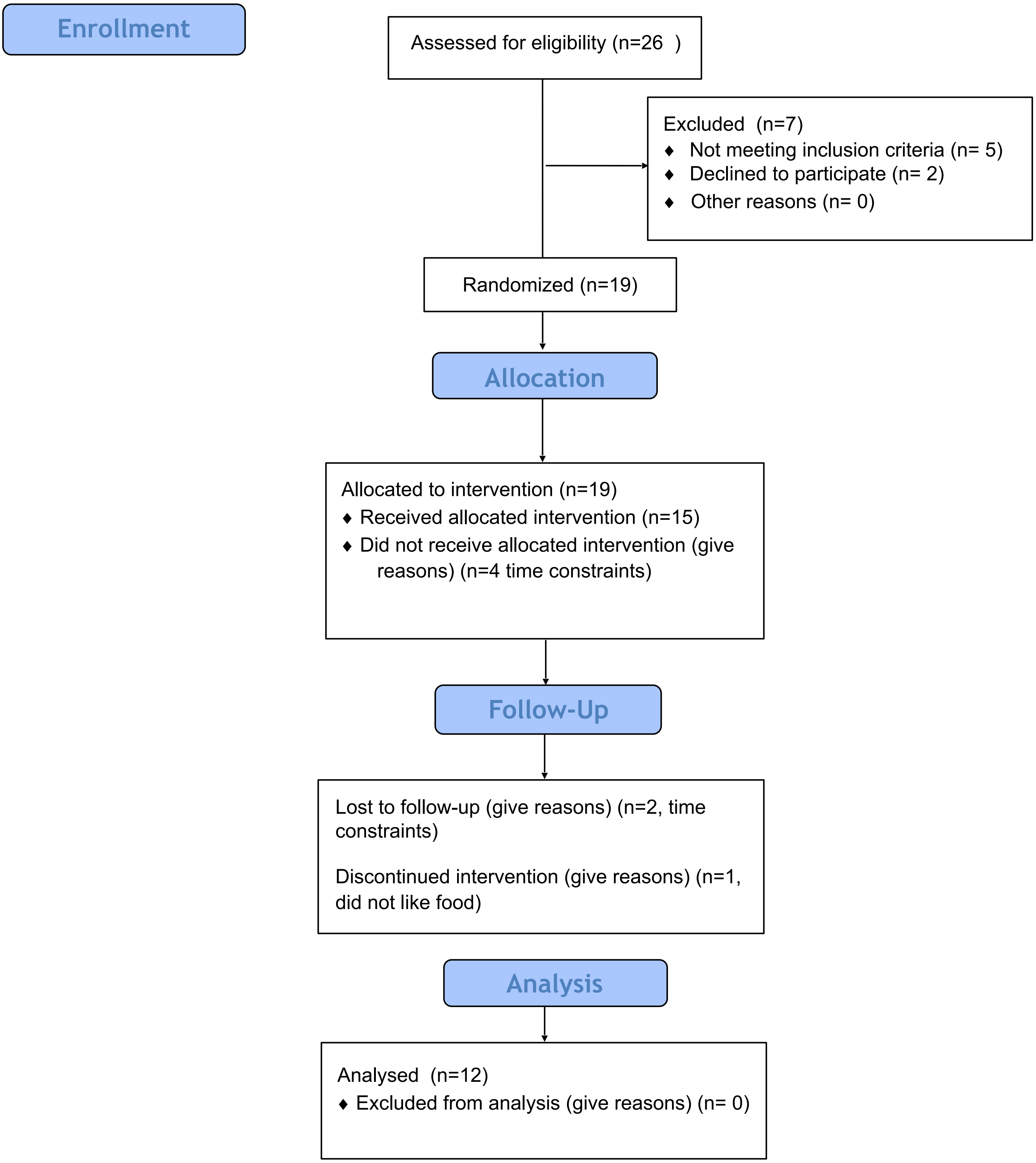


Figure S1: CONSORT flow diagram of study

| | | Dose of carbohydrate | | | | |
|---|---|---|---|---|---|---|
| | | 10% | 15% | 20% | 25% | 30% |
| | | g | g | g | g | g |
| Breakfast | Bagels | 20 | 35 | 50 | 60 | 80 |
| | Butter | 22 | 24 | 18 | 15 | 15 |
| | Keto Pancake | 125 | 120 | 105 | 100 | 90 |
| | Walnuts | 25 | 20 | 25 | 25 | 20 |
| Lunch | White fish | 80 | 80 | 80 | 70 | 60 |
| | Rice | 35 | 65 | 100 | 130 | 150 |
| | Cauliflower | 60 | 60 | 60 | 60 | 60 |
| | Sunflower oil | 22 | 22 | 22 | 22 | 17 |
| | Lettuce | 35 | 35 | 35 | 35 | 35 |
| | Cucumber | 50 | 50 | 50 | 50 | 50 |
| | Tomatoes | 50 | 50 | 50 | 50 | 50 |
| | Feta cheese | 60 | 60 | 54 | 55 | 60 |
| | Olive oil | 25 | 20 | 19 | 15 | 15 |
| Dinner | Chicken breast | 60 | 60 | 55 | 50 | 50 |
| | Croutons | 20 | 30 | 40 | 50 | 60 |
| | Avocado | 130 | 120 | 110 | 100 | 70 |
| | Parmesan cheese | 15 | 15 | 12 | 15 | 15 |
| | Lettuce | 30 | 30 | 30 | 30 | 30 |
| | Olives | 18 | 18 | 18 | 18 | 18 |
| | Olive oil | 25 | 22 | 20 | 17 | 15 |
| | Dates | 10 | 15 | 20 | 25 | 40 |
| Snack | Greek yoghurt | 120 | 160 | 140 | 150 | 150 |
| | Cream | 10 | 10 | 5 | 0 | 0 |
| | Raspberries | 10 | 25 | 30 | 35 | 40 |

| | | | | | | |
|---|---|---|---|---|---|---|
| | Mango | 0 | 30 | 50 | 90 | 115 |
| | Pistachio nuts | 25 | 10 | 17 | 15 | 12 |

Table S1: Example of how the menu is altered with each dose.

| | **Mean 24-hour** | **Post-prandial** | **CONGA** | **MAGE** | **log(LBGI)** | **HBGI** |
|---|---|---|---|---|---|---|
| Constant | 16.949*** (3.809) | 18.217*** (4.603) | 4.294* (1.952) | 11.285* (5.215) | -12.029* (5.366) | 19.721* (7.579) |
| Dose | 0.017 (0.009) | 0.021 (0.011) | 0.007 (0.008) | 0.014 (0.019) | -0.024 (0.029) | 0.032 (0.022) |
| Age | -0.166* (0.069) | -0.173 (0.083) | -0.047 (0.035) | -0.128 (0.094) | 0.179 (0.095) | -0.310 (0.136) |
| Male | -2.205* (0.719) | -2.717* (0.869) | -0.662 (0.364) | -1.836 (0.976) | 3.007* (0.988) | -3.514* (1.426) |
| Kuwati | 0.963 (0.715) | 0.674 (0.864) | -0.072 (0.365) | -0.263 (0.977) | -1.967 (1.000) | 2.447 (1.421) |
| Weight change (kg) | 0.076 (0.098) | 0.118 (0.120) | | 0.474* (0.195) | 0.219 (0.295) | 0.326 (0.233) |
| Mean steps per day | -0.049 (0.044) | (0.054) | 0.007 (0.033) | 0.024 (0.082) | -0.033 (0.109) | -0.023 (0.102) |
| % Variance explained† | 70% | 69% | 42% | 49% | 22% | 60% |

Table S3: Parameter estimates with standard errors from the linear mixed-effects models (*p<0.05; **p<0.01; ***p<0.001)

†Random effect for intercept explains xx% of the variance that remains after the variance has been explained by the fixed effects.

Supplementary data: Figure S2: Post-prandial glucose excursions for each participant per meal, per day.

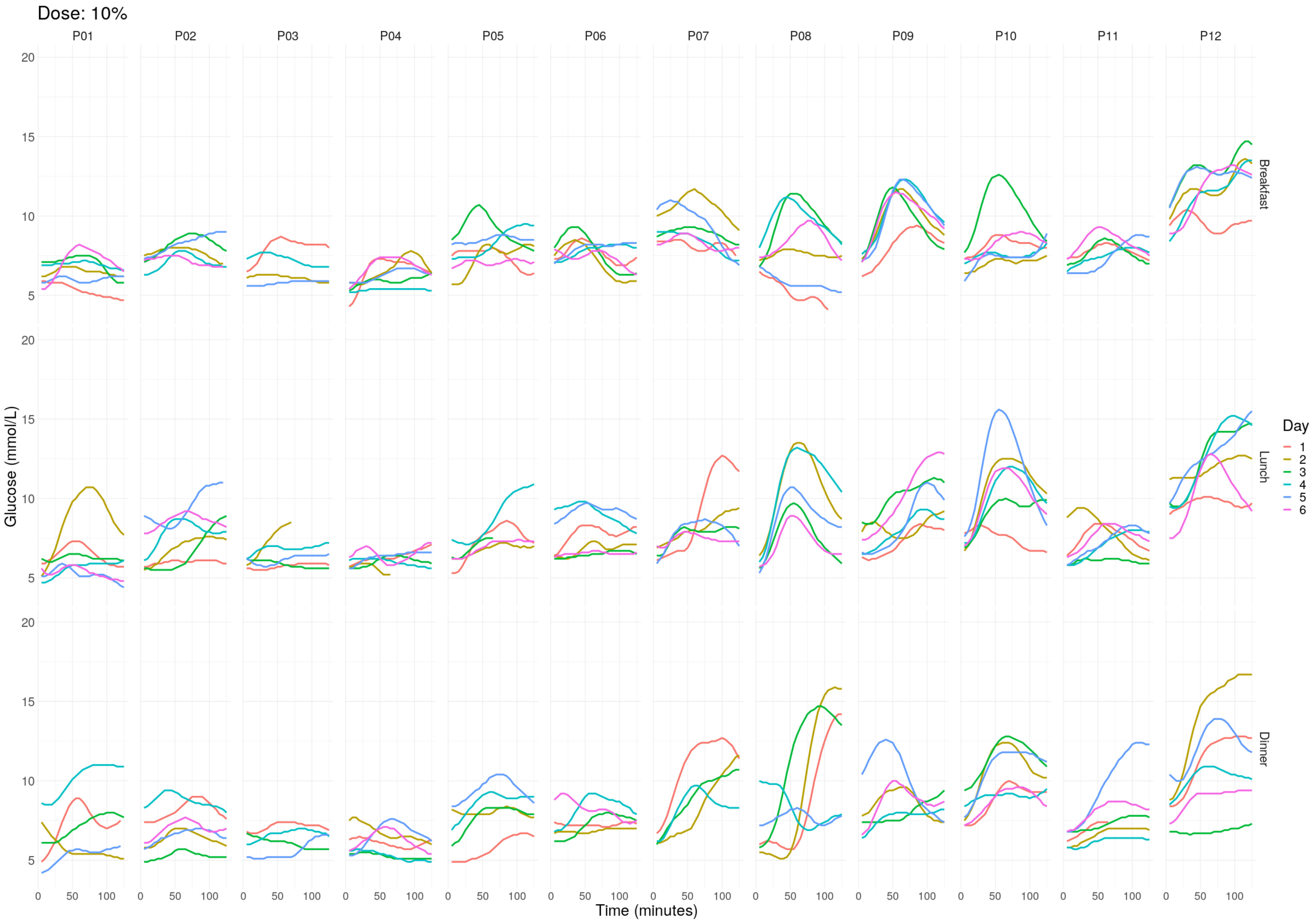

Supplementary data: Figure S2: Post-prandial glucose excursions for each participant per meal, per day.

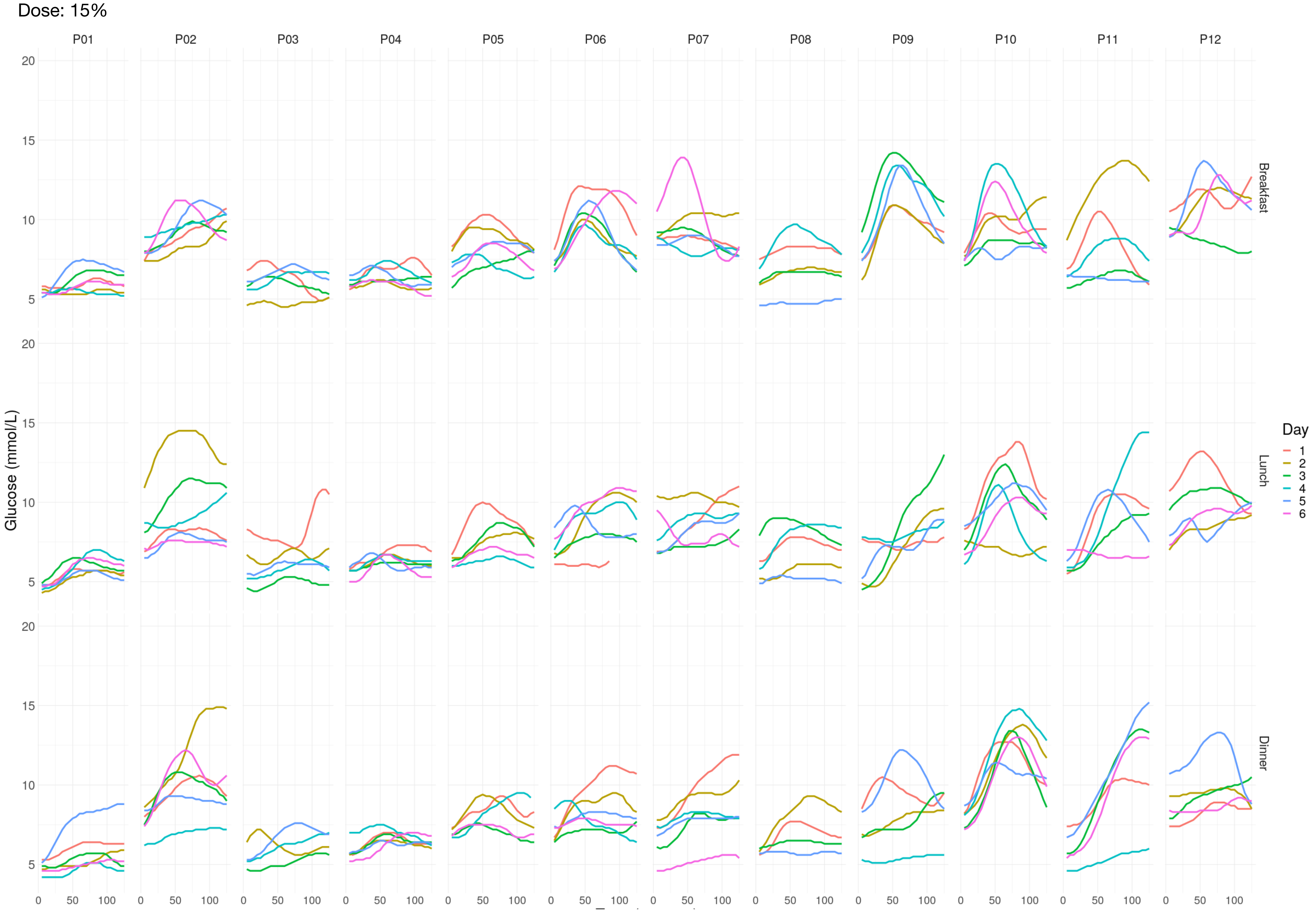

Supplementary data: Figure S2: Post-prandial glucose excursions for each participant per meal, per day.

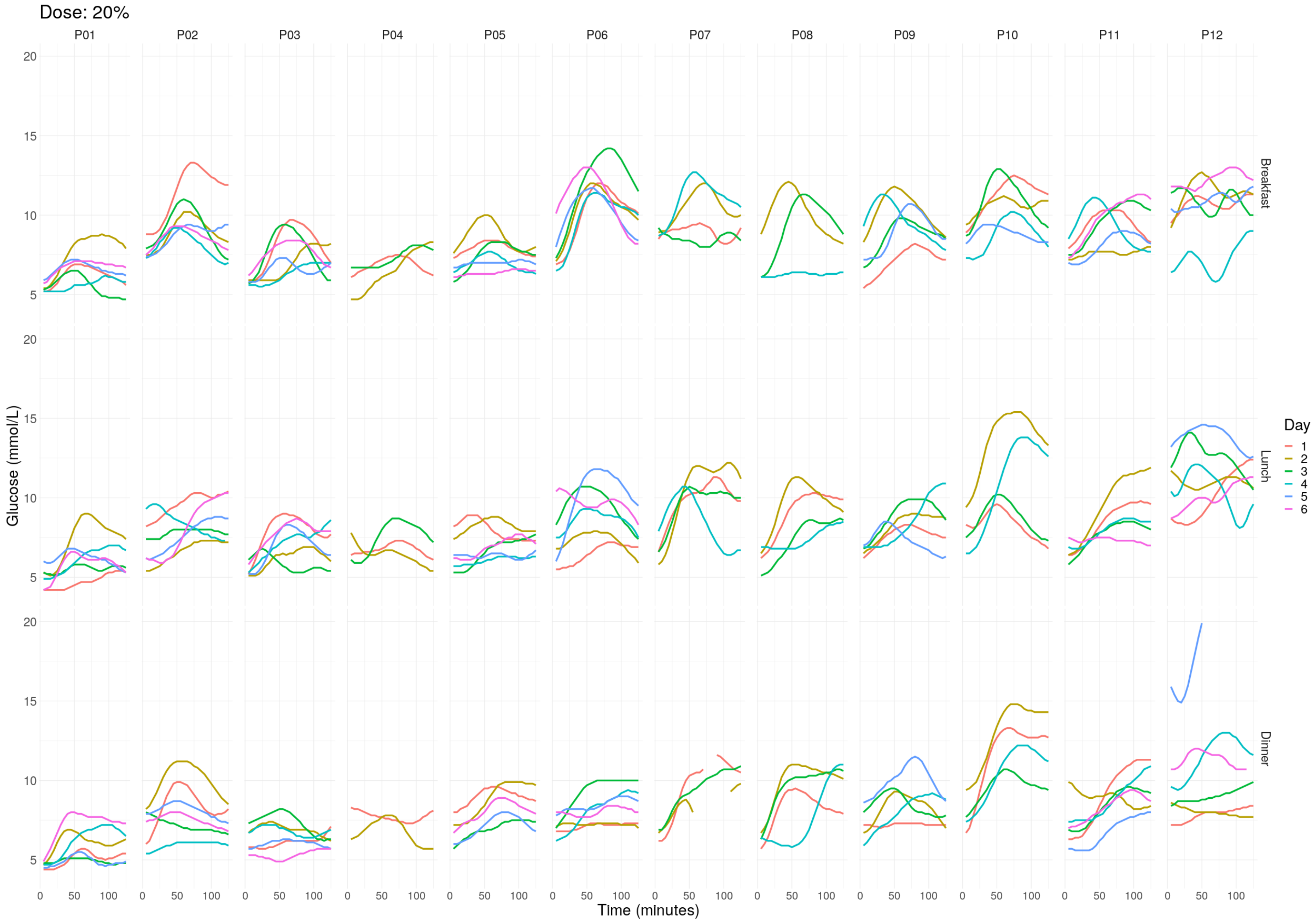

Supplementary data: Figure S2: Post-prandial glucose excursions for each participant per meal, per day.

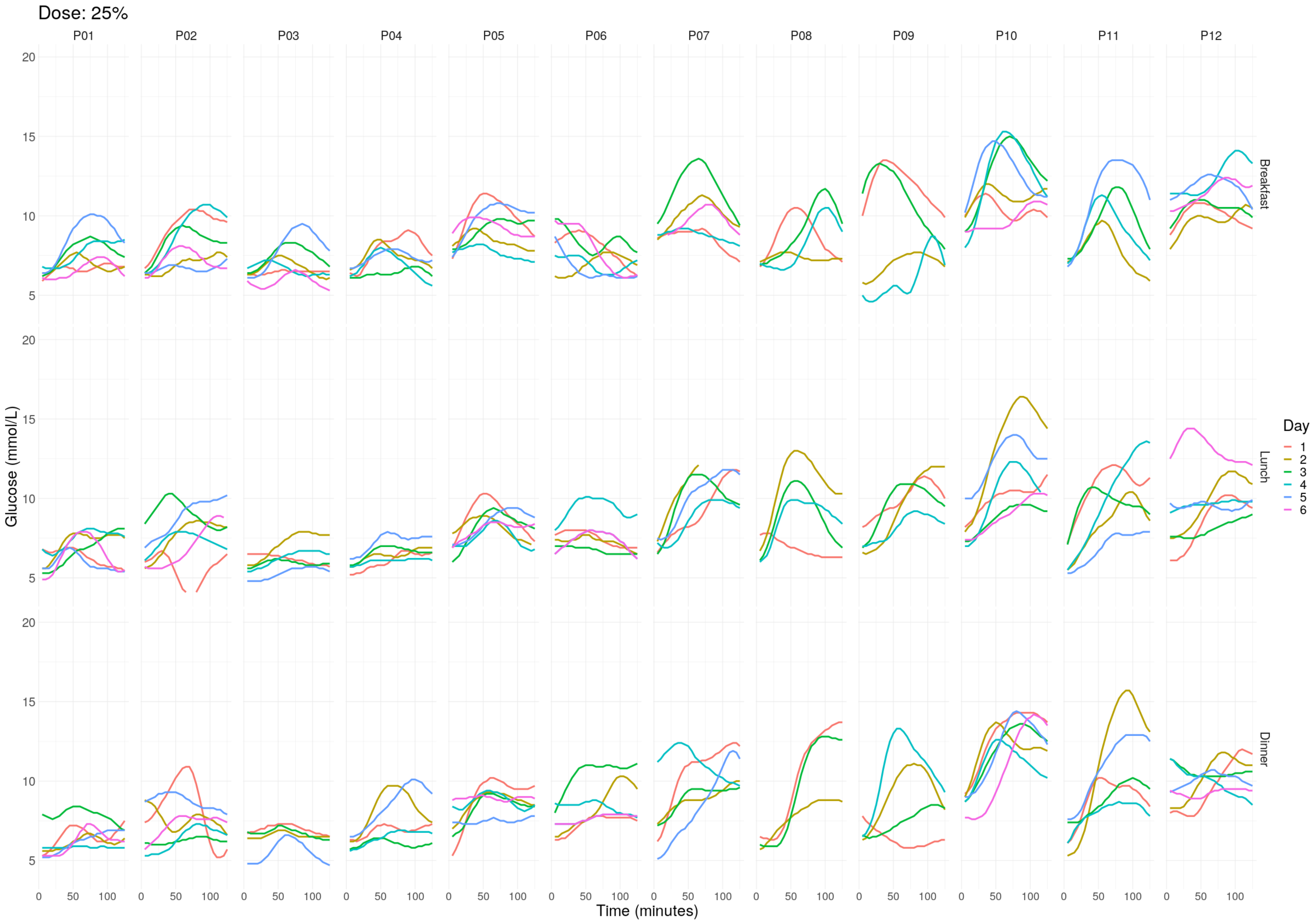

Supplementary data: Figure S2: Post-prandial glucose excursions for each participant per meal, per day.

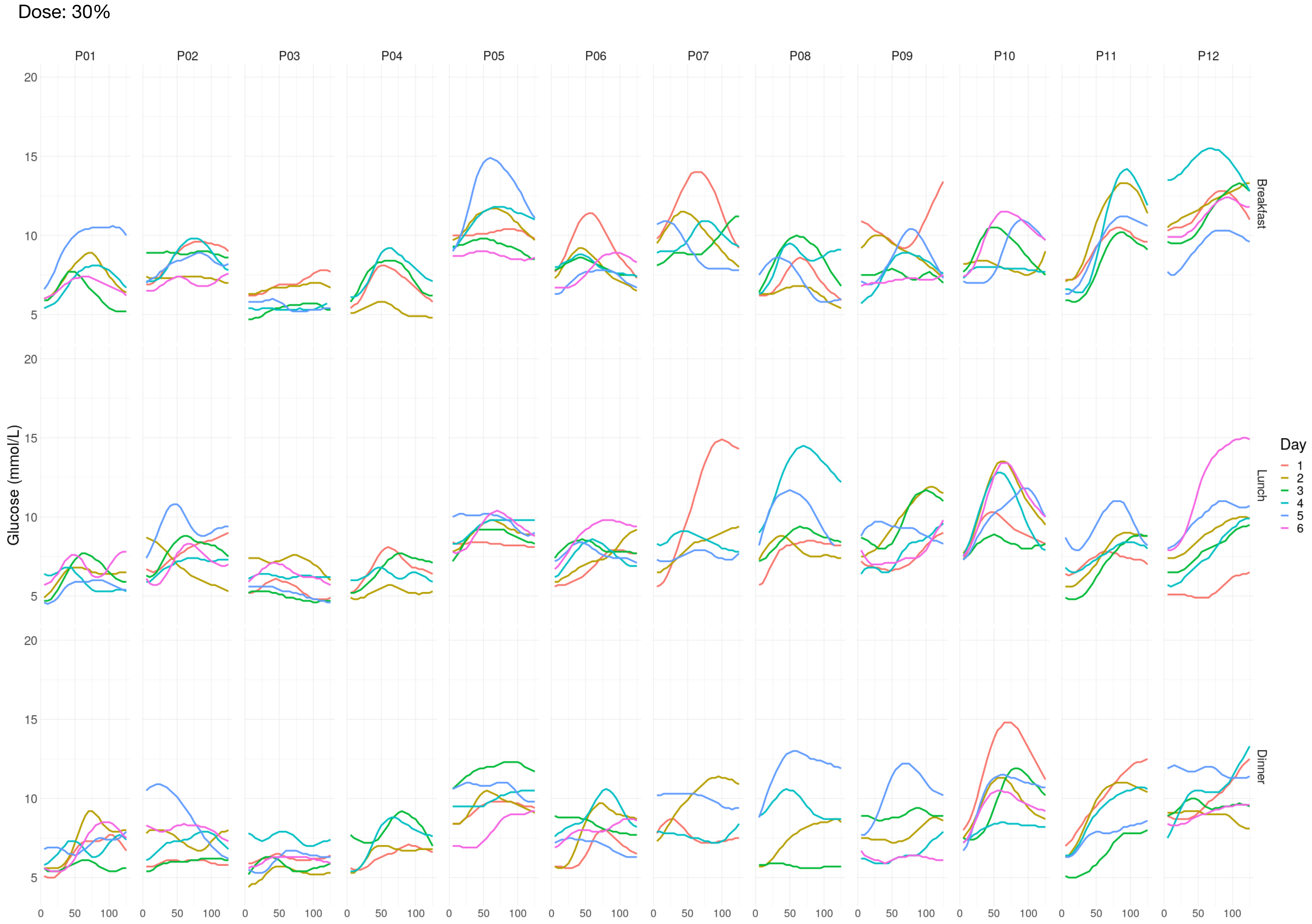
